# Pitfalls and solutions in case fatality risk estimation – A multi-country analysis on the role of demographics, surveillance, time lags between reporting and death and healthcare system capacity on COVID-19

**DOI:** 10.1101/2020.05.16.20104117

**Authors:** Patrizio Vanella, Christian Wiessner, Anja Holz, Gérard Krause, Annika Möhl, Sarah Wiegel, Berit Lange, Heiko Becher

## Abstract

European countries report large differences in COVID-19 case fatality risk (CFR) and high variation over the year. CFR estimates may both depend on the method used for estimation and of country-specific characteristics. While crude methods simply use cumulative total numbers of cases and deaths, the CFR can be influenced by the demographic characteristics of the cases, case detection rates, time lags between reporting of infections and deaths and infrastructural characteristics, such as healthcare capacities.

We used publicly available weekly data from the national health authorities of Germany, Italy, France and Spain on case and death numbers by age group connected to COVID-19 for the year 2020. We propose to use smoothed data of national weekly test rates for case adjustment and investigated the impact of different time lags from case reporting to death on the estimation of the CFR. Finally, we described the association between case fatality and the demand for hospital beds for COVID-19, taking into account national hospital bed capacities.

Crude CFR estimates differ considerably between the four study countries with end-of-year values of approximately 1.9%, 3.5%, 2.5% and 2.7% for Germany, Italy, France and Spain, respectively. Age-adjustment reduces the differences considerably, resulting in values of 1.61%, 2.4% and 2% for Germany, Italy and Spain, respectively. France’s age-specific data was restricted to hospitalised cases only and is therefore not comparable in that regard. International crude International CFR time series show smaller differences when adjusting for demographics of the cases or the test rates. Curves adjusted for age structure, testing or time lags show smaller variance over the year and a smaller degree of non-stationarity. The data does not suggest any connection of CFRs to hospital capacities for the four countries under study.

## Introduction

Countries have reported divergent case fatality estimates of COVID-19. The crude case fatality risk (CFR) estimate, namely the cumulative number of deaths divided by the cumulative number of cases, is known to be biased (Lipsitch et al. 2015). The main sources of bias are given in Table 1. A distinction must be made between factors that might influence the actual lethality (as denoted by ^*^), such as healthcare capacity and those that bias the estimates of the CFR, such as an underassessment of cases.

**Table 1.**
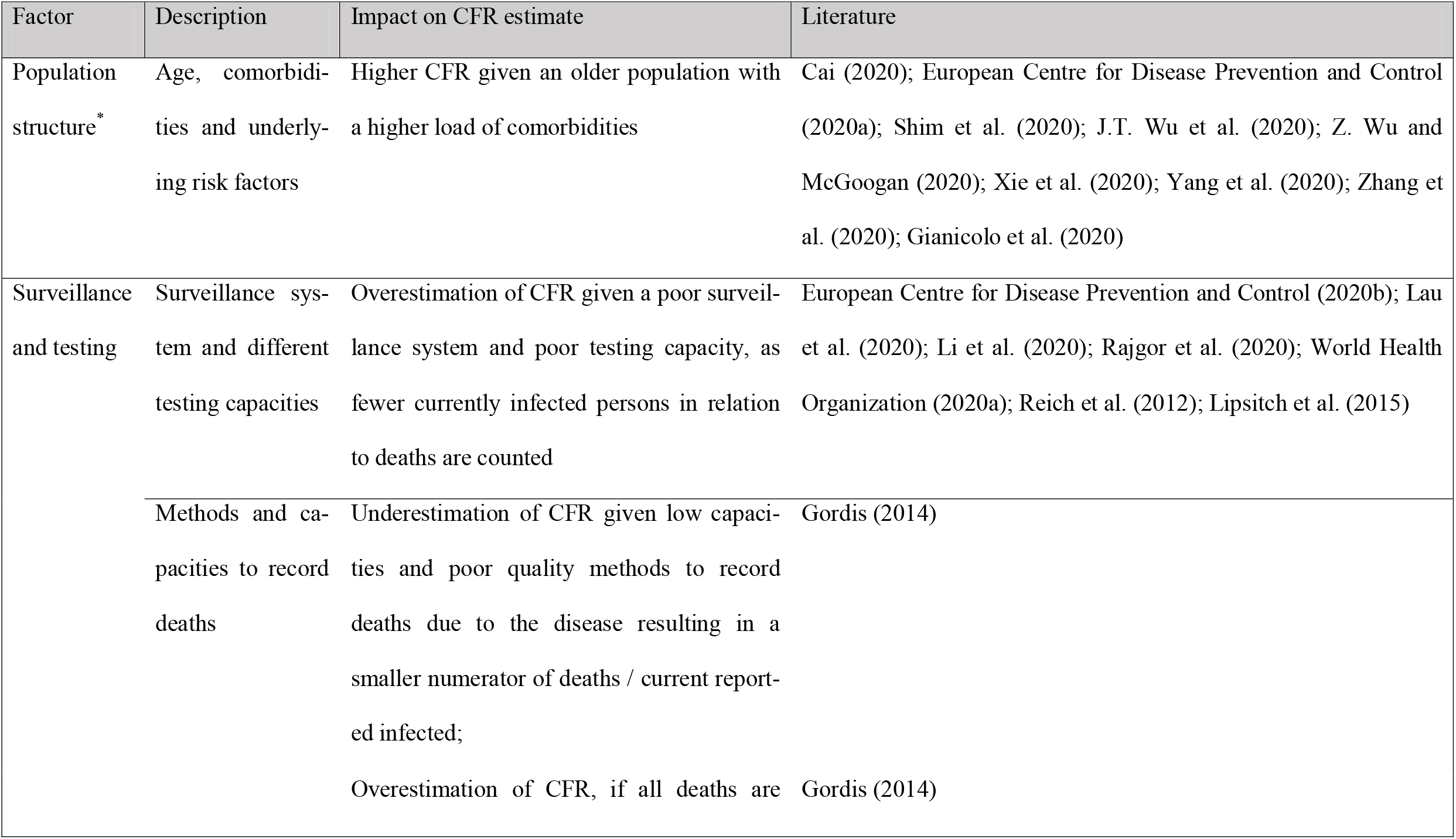
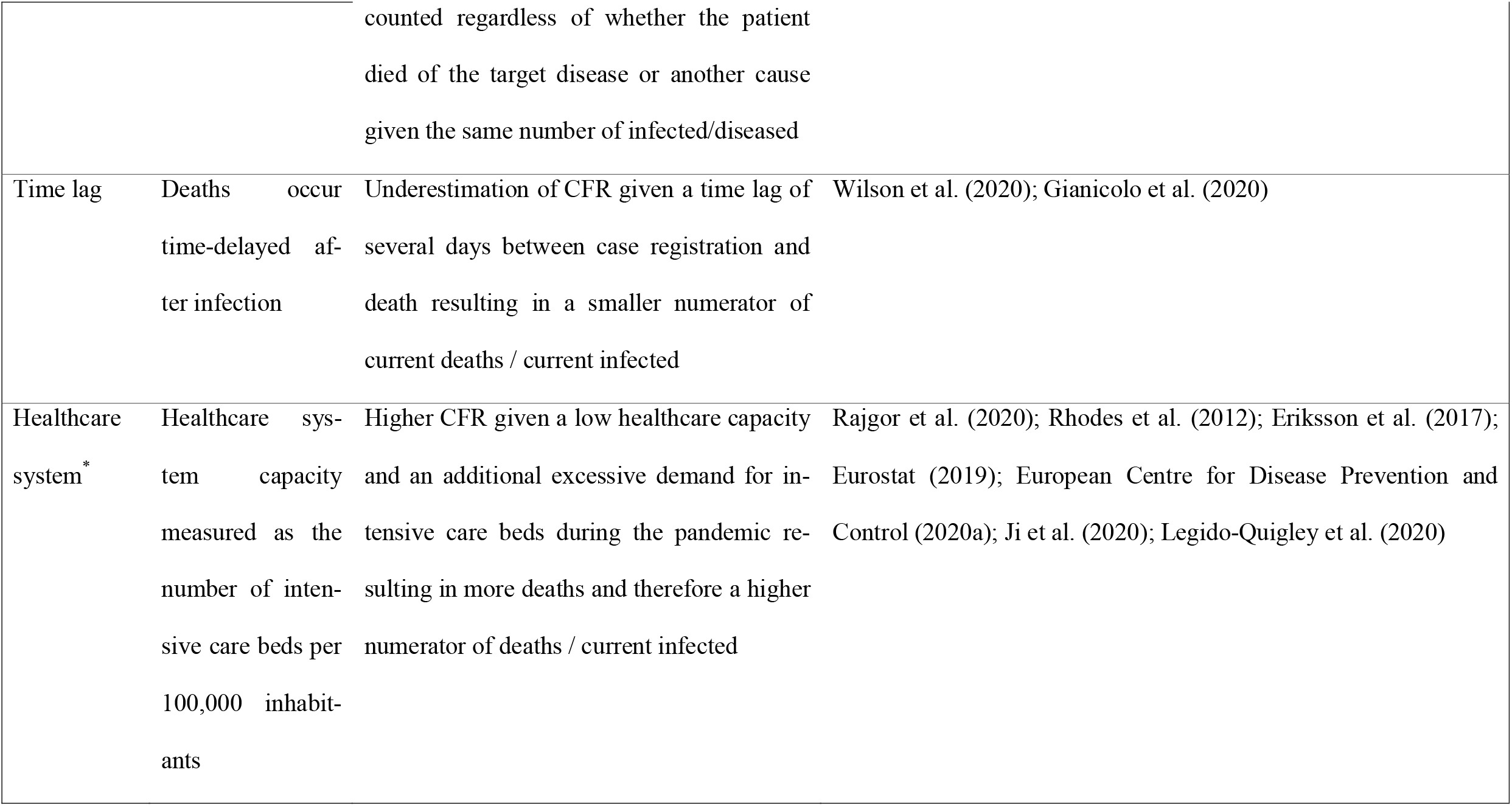
Sources of bias of case fatality risk estimates

Different demographic characteristics of infected cases regarding age, comorbidities or underlying risk factors, as well as different underlying population structures of the respective countries, might explain different CFRs (Dudel et al. 2020). There is evidence that old-age and comorbidities – such as hypertension, diabetes, cardiovascular disease or chronic lung disease – are major risk factors for severe COVID-19 infection outcomes (Zhou et al. 2020; Jordan, Adab, and Cheng 2020; J.T. Wu et al. 2020). The higher susceptibility to disease as well as the higher prevalence of comorbidities in the elderly has an impact on the morbidity and mortality of this subpopulation resulting in higher CFRs in countries with an older population compared to countries with a younger age structure, which is apparent in the COVID-19 pandemic (J.T. Wu et al. 2020; Z. Wu and McGoogan 2020; Shim et al. 2020; European Centre for Disease Prevention and Control 2020a; Cai 2020; Xie et al. 2020; Yang et al. 2020; Zhang et al. 2020; Dudel et al. 2020). Different surveillance systems and different testing capacities across countries lead to huge variations in the number of tests performed (European Centre for Disease Prevention and Control 2020b; World Health Organization 2020b; Rajgor et al. 2020; Pan et al. 2020; Fang et al. 2020; European Centre for Disease Prevention and Control 2021b). Underassessment of reported infections therefore differs among countries (Li et al. 2020; Lau et al. 2020). Furthermore, surveillance and testing capacities influence the probability of detecting infections early and thus enact countermeasures. Capacities and methods to record deaths as having been caused by COVID-19 may also differ between countries (Gordis 2014). While in some countries, post-mortem screening of all deaths has been installed, other countries are only performing this when there is clinical suspicion (Onder, Rezza, and Brusaferro 2020). Moreover, during the pandemic, the countries have changed their testing strategy and number of tests performed multiple times (Robert Koch Institut 2020a, 2020b; European Centre for Disease Prevention and Control 2021b), limiting the representativeness of case time series on the country level.

There exists a time lag between reporting of an infection and the eventual death of said individual. The distribution of the time lag may differ between countries. This delay is not reflected in crude CFR estimates (Wilson et al. 2020). A more robust estimate would be given by dividing cumulative deaths by cumulative recoveries. This, however, is not a reliable estimate as well due to a low number of recoveries during the early stages of the pandemic, when a high relative increase of infection numbers and an incomplete reporting of recoveries are witnessed (Lipsitch 2020). Therefore, some authors propose to investigate the cumulative deaths in relation to lags of various days of the cumulative infection numbers (Wilson et al. 2020; Lipsitch 2020). However, as a result of the high transmission rates of the virus in the early stages of an epidemic, the estimates depend strongly on the appropriate lag and both under- and overestimation of the true CFR can occur (Spychalski, Blazynska-Spychalska, and Kobiela 2020).

Furthermore, CFRs may be influenced by the healthcare system capacity of the affected countries. Previous studies have shown that healthcare capacities differ substantially among countries and even among regions within countries (European Centre for Disease Prevention and Control 2020a; Rhodes et al. 2012; Eurostat 2019; Legido-Quigley et al. 2020; Eriksson et al. 2017; Ji et al. 2020; Rajgor et al. 2020; OECD 2021a). A healthcare system overwhelmed by the pandemic may be associated with higher CFRs.

All factors mentioned above may explain differences in CFRs in affected countries to a certain degree at different time points during the COVID-19 pandemic. It is unclear, how much of the difference in during-epidemic CFR estimates is explained by each of these factors. This paper aims at quantifying the effect of demographics, the level of testing, delays in death after infection and demand for hospital beds on weekly CFR estimates of COVID-19 in Germany, Italy, France and Spain during the year 2020 in a comparative perspective. The selected countries are interesting for our study, as they are the four most populous countries in the European Union (EU), covering more than half of the total population of the EU (Eurostat 2020). Moreover, these countries show different levels in the COVID-19 CFR time series and have borne a large part of the COVID-19 disease burden over the study period.

## Data and methods

We obtained weekly numbers of cases and deaths of Germany (Robert Koch Institut 2021), Italy (Istituto Superiore di Sanità 2020a, 2020b, 2020c, 2020d), France (Santé publique France 2020a, 2020b) and Spain (Instituto de Salud Carlos III 2020a, 2020b) by age group for the year 2020 as provided from the websites of the national health authorities. For comparison of crude and age-adjusted CFRs, we use the European Standard Population (Gesundheitsberichtserstattung des Bundes 2020) for standardisation. The impact of surveillance of cases is integrated by considering weekly time series data of per 100,000 inhabitant test rates, as provided by the European Centre for Disease Prevention and Control (ECDC) (2021b). The effect of different time lags on CFR estimates is analyzed using data provided daily by Johns Hopkins University (JHU) (2021). For our investigation of the impact of the healthcare system capacity, we use estimates for the available hospital beds per 100,000 inhabitants by the OECD (2021a) and counts of weekly new hospitalisations due to COVID-19, reported regularly by the ECDC (European Centre for Disease Prevention and Control 2021a).

We use the following notation: 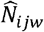 and *D*_*ijw*_ denote the total cumulative number of cases and deaths, respectively, for age group *i*, country *j* and up to week *w. d*_*.jk*_ denotes the number of deaths during week *k*, hence 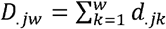. Similarly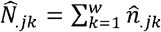, with 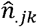 denoting the number of observed new cases over all age groups in country *j* during week *k*. The true number of new infections in age group *i*, country *j* and week *k* (denoted by *n*_*ijk*_), is latent (unknown). For the analysis, we use either the observed number 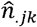 or the number adjusted for country-specific test capacity, *ñ*_*ijk*_. The number of cumulative infections in said group up to week *w* is 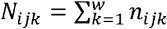.

With this notation, the sequence of our analysis is as follows. The crude CFR, which ignores all of the factors mentioned earlier, in week *w*, in country *j* is:

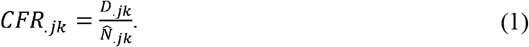

Figure 1 illustrates the development of the crude CFR estimates of the four selected countries in percent between March 4^th^ (i.e. calendar week 10) and December 30^th^ (i.e. calendar week 53), 2020.

**Figure 1.**
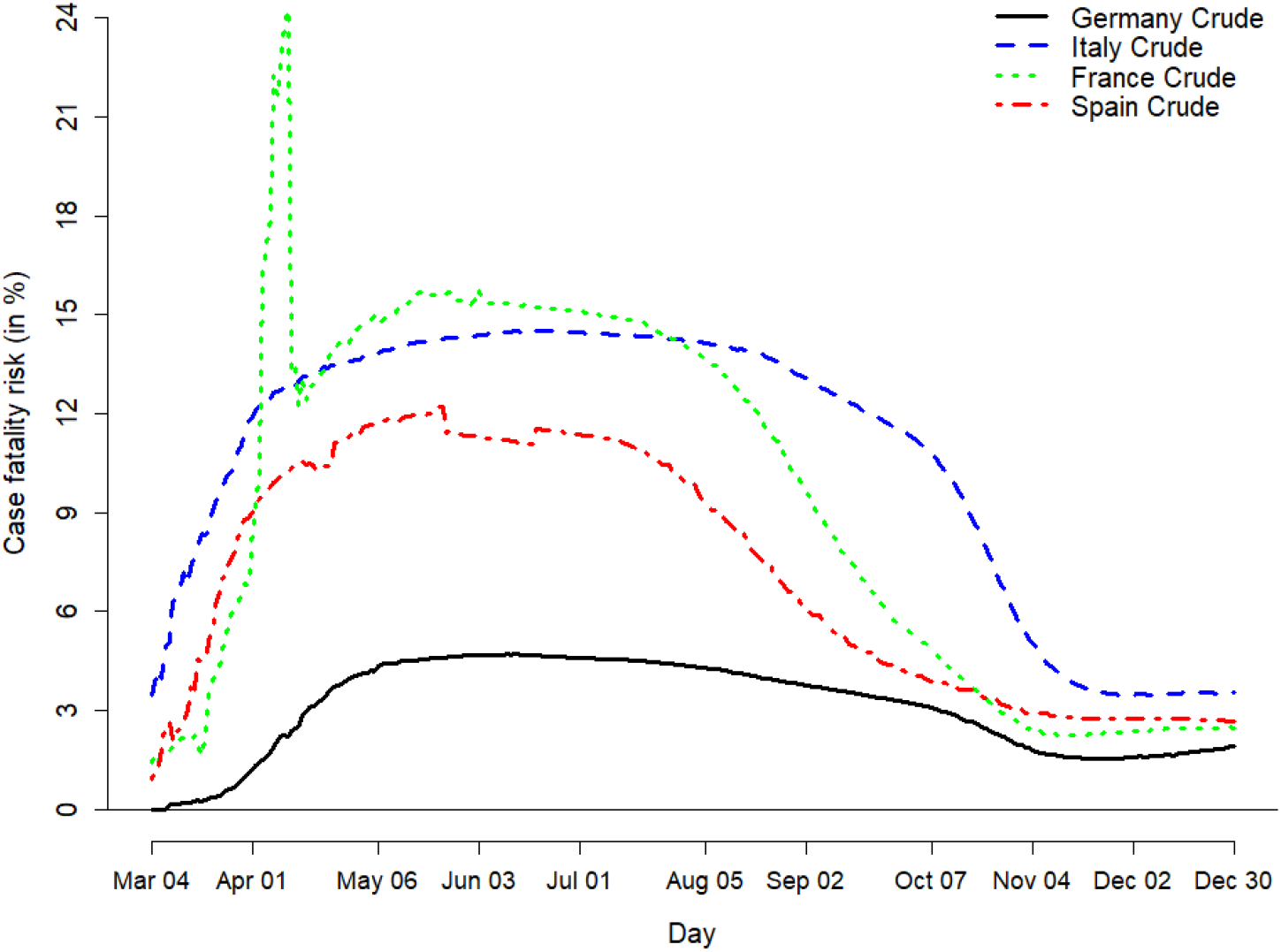
Crude case fatality risk estimates due to COVID-19 between March 4^th^ and December 30^th^, 2020. Sources: Johns Hopkins University (2021); Own computation and design

All curves increased until late spring / early summer 2020, before decreasing again until the end of the study period. We observed significant differences between the curves. Our study aims to explain these differences to some extent and develop adjusted case fatality measures. For the French and the Spanish line, a quite jagged course is noticeable. Especially a sharp increase until early April for France can be observed, followed by a sharp decrease on April 12^th^. This is due to an almost doubling of the case numbers in the JHU data on that date following the addition of French Ehpad data on cases reporting in nursing homes (Johns Hopkins University 2021; Ministère des Solidarités et de la Santé 2021). Therefore, we have a significant undercounting of cases in France before that date resulting in overestimation of CFR estimates during that period. The tub shape in Spain observed between mid-May and mid-June could be explained by a change in reporting of the Spanish COVID-19 data during that time. Between mid-May and early July, Spanish authorities developed a new strategy in tracking and reporting of COVID-19 data (Instituto de Salud Carlos III 2020b), resulting in a lack of detailed reports in the meantime, which could lead to unrepresentative case data during that time. By the end of the year, the crude CFR estimates have converged somehow, but there are still differences between Germany, Italy and France / Spain.

Our first aim is to identify the role of the cases’ age structure in the overall CFRs and to derive age-specific and age-standardised CFR estimates of Germany, Italy, France and Spain. Based on cumulative age-specific case numbers *N*_*ijk*_ of the four European countries, we calculated age-specific CFR estimates

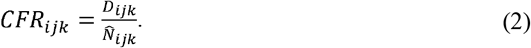

Multiplying these with population weights derived from the European Standard Population, we receive age-standardised CFR estimates

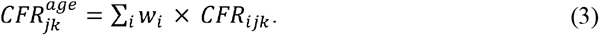

For the second aim, namely, to investigate the effect of testing on the CFR estimates, we use data on weekly per 100,000 person test rates provided by ECDC (European Centre for Disease Prevention and Control 2021b). Our baseline assumption is that all infections have been discovered in the calendar week, where the global maximum of country-specific tests has been performed. Limitations of this assumption are given in the discussion. We then assume that the weekly share of detected cases is proportional to the ratio of the observed case number and the number of tests per 100,000 performed, say for a certain week *k*:

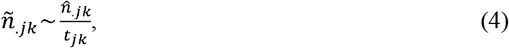

with *ñ*_.*jk*_ being the test-adjusted number of cases in country *j* during week *k* and 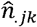 being the corresponding number of observed cases. *t*_jk_ denotes the number of tests performed per 100,000 in week *k* for country *j*. Since the number of tests is not given by age group, we assume a mean detection rate over all ages. Furthermore, since the reported numbers vary considerably between weeks, we calculate a moving average with weights 0.25, 0.5 and 0.25 for weeks *k* − 1, k and *k* + 1, respectively, to obtain the estimate for week *k*. It follows that

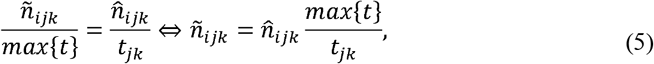

with *max*{*t*} denoting the maximum of weekly tests performed in the study countries over the study period.

For *max*{*t*}, we obtain 3568.70 per 100,000, the filtered value for France in calendar week 52, 2020. Replacing the observed number of cases 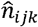 with the adjusted estimated number *ñ*_*ijk*_ yields the test-adjusted cumulative case numbers 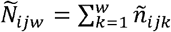. We then adjust the estimates from (1) – (3) to test capacity accordingly to derive test-adjusted crude and age-specific CFR estimates.

The crude test-adjusted CFR until week *w* in country *j* is then

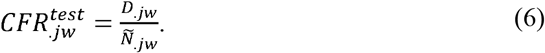

Accordingly, this becomes

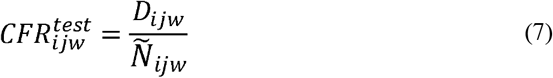

for age group *i*, which is a test-adjusted CFR for said age group.

For Aim 3, the investigation of different lags on the CFR estimate, the unknown distribution of the time lag _Δ_ between reporting of a case and death is considered. Verity et al. (2020) estimated the average time from infection to death to be about 14 days. Thus, the average time from reporting a case to death is several days shorter. The lag-adjusted CFR estimate without age- or test-adjustment is given by

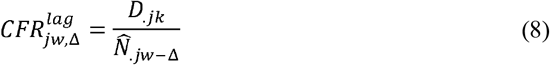

for the pandemic over the study period with *Δ 0, 1, 2, 3*.

We also investigate a weighted average as

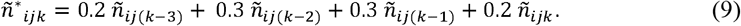

The fourth aim is to investigate the association between the CFR and the healthcare capacity of a specific country. As a suitable measure, we suggest the number of COVID-19 related fatalities during a particular week divided by the number of hospital admissions due to COVID-19 of the previous *γ* weeks, *h*_.*jk,γ*_. We will first of all give a qualitative assessment of this connection by graphical analysis. To account for the national hospital bed capacities in a comparative international analysis, we adjust the weekly new hospitalisations, as provided by ECDC (European Centre for Disease Prevention and Control 2021a), by the bed capacities per 1,000 inhabitants. These are provided as a static statistic by the OECD (2021b). As both measures are measured relative to the country’s population, dividing them leads to an admission-per-hospital bed measure in percent, defined as

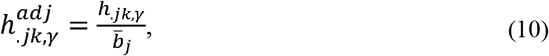

with 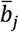 denoting the per 100,000 inhabitant number of available hospital beds in country *j*.

Whereas this measure has its merits, as it is static and hence does not change over time, it should serve as a rough adjustment parameter, which accounts for the differences among national

We present the results of our country-specific analyses in the next section.

## Results

### Aim 1. During-epidemic crude and age-standardised CFR estimates, alongside CFRs of Germany, Italy, France and Spain

Figure 2 illustrates weekly estimates of crude and age-standardised CFRs against the crude CFR time series from JHU, estimated as explained in the section Data and Methods. More detailed results can be found in the online supplement.

**Figure 2.**
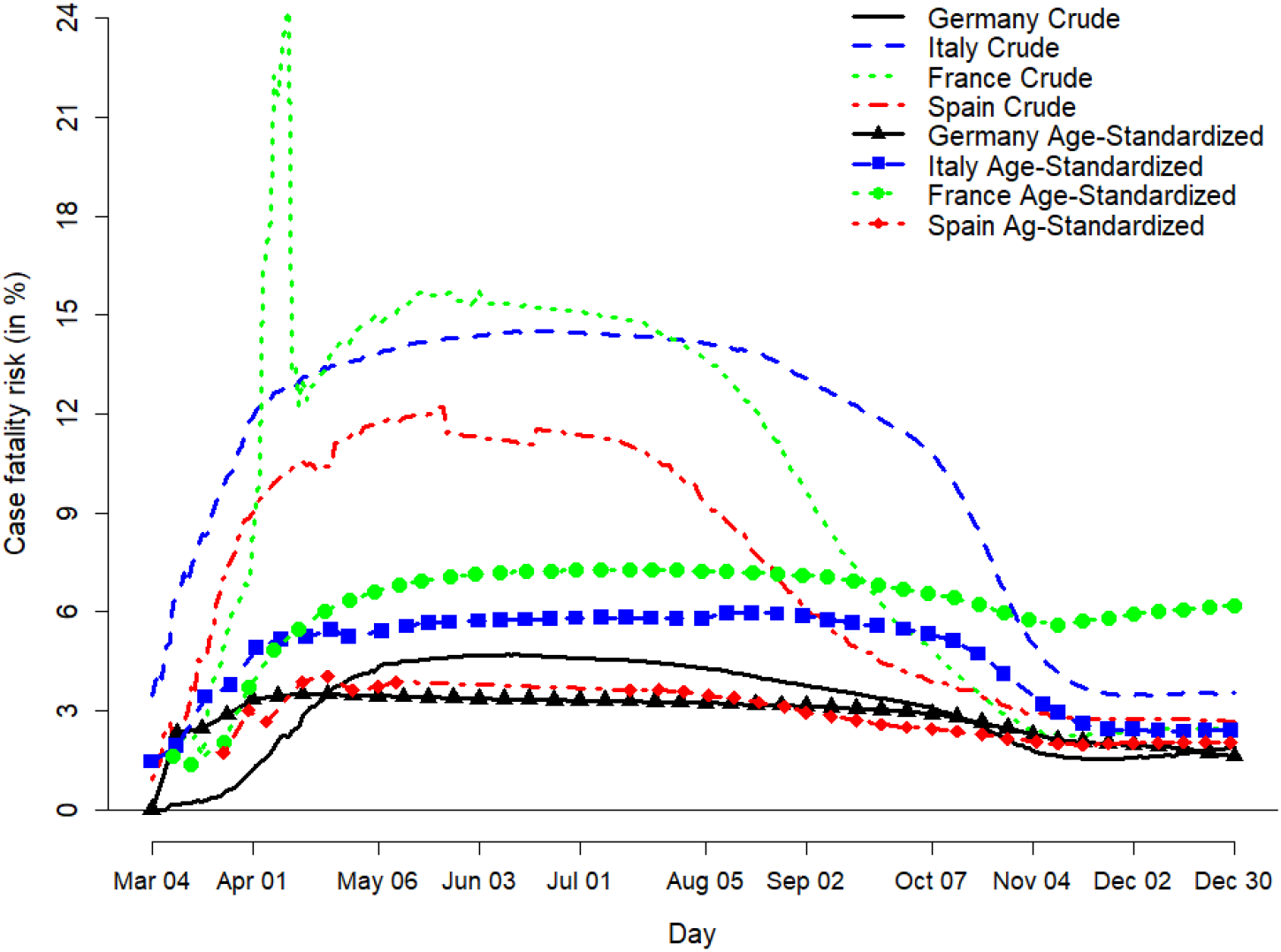
Crude compared to age-standardised CFR estimates. Sources: Gesundheitsberichtserstattung des Bundes (2020); Istituto Superiore di Sanità (2020a, 2020b, 2020c, 2020d); Santé publique France (2020a, 2020b); Robert Koch Institut (2021); Instituto de Salud Carlos III (2020a, 2020b); Johns Hopkins University (2021); own computation and design

The courses of the age-standardised curves are more stable than the crude CFR curves. A large share of the decreases observed in crude CFRs since Spring 2020 vanishes when accounting for the age structure of the cases. In general, the weekly investigation smoothes out some of the variations we observe in daily monitoring.

### Aim 2. Account for bias due to different levels of testing over time and among countries

Figure 3 shows the test-adjusted CFRs computed according to (7) for the four study countries in comparison to the crude CFRs. We see that adjustment of the case numbers to testing activity already reduces the non-stationarity in the crude CFR significantly, even if not accounting for the other factors influencing the CFR estimates. This is especially apparent in the CFR series of Germany, Italy and France, where the adjusted curves have been relatively stable since the beginning of May 2020, indicating a large bias in the crude CFR estimates over time. We especially see that the decreasing trends in the CFR series since summer are heavily affected by increased testing.

**Figure 3.**
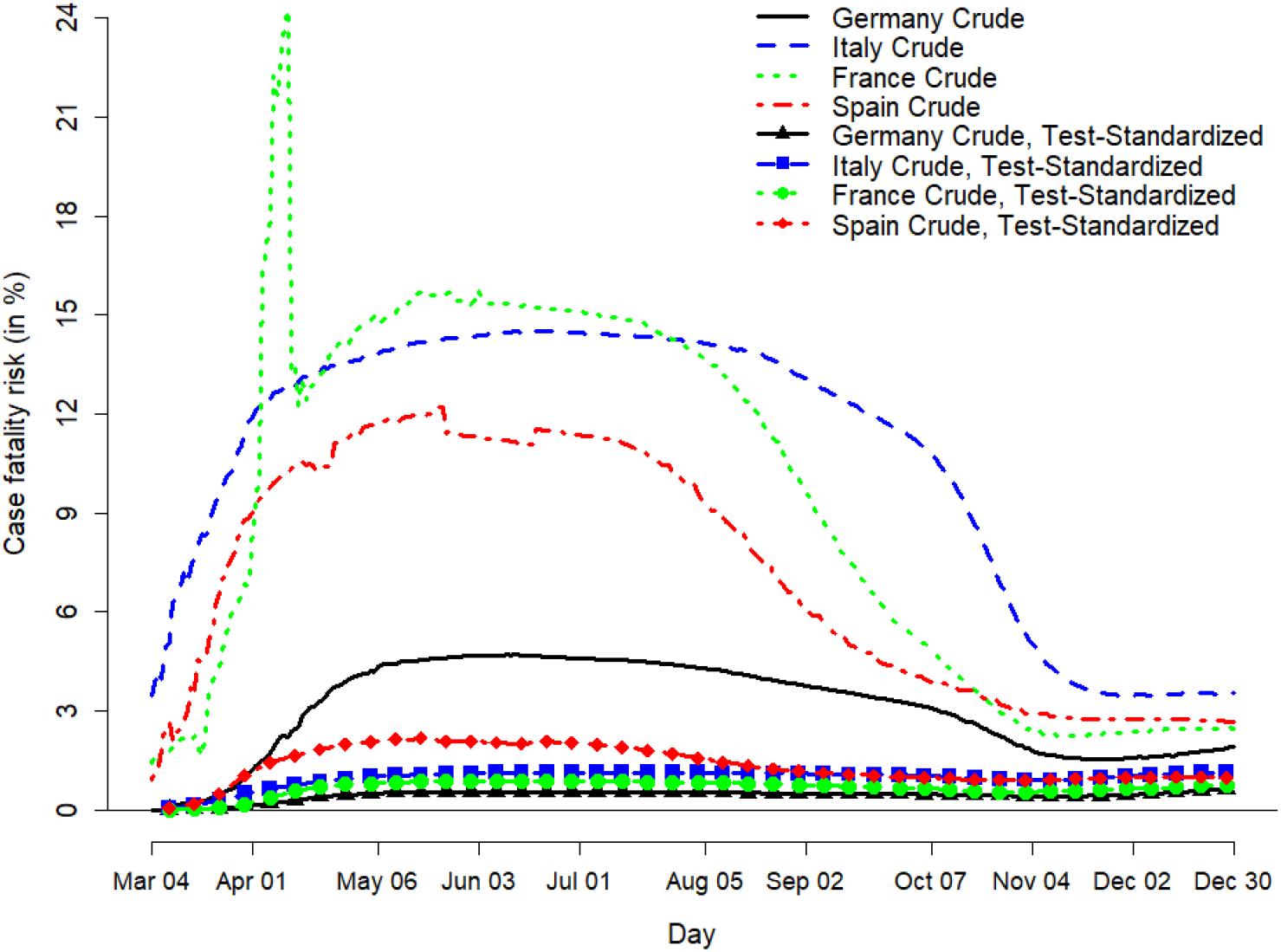
Crude versus test-standardised CFRs. Sources: Johns Hopkins University (2021); European Centre for Disease Prevention and Control (2021b); Own computation and design

**Figure 4.**
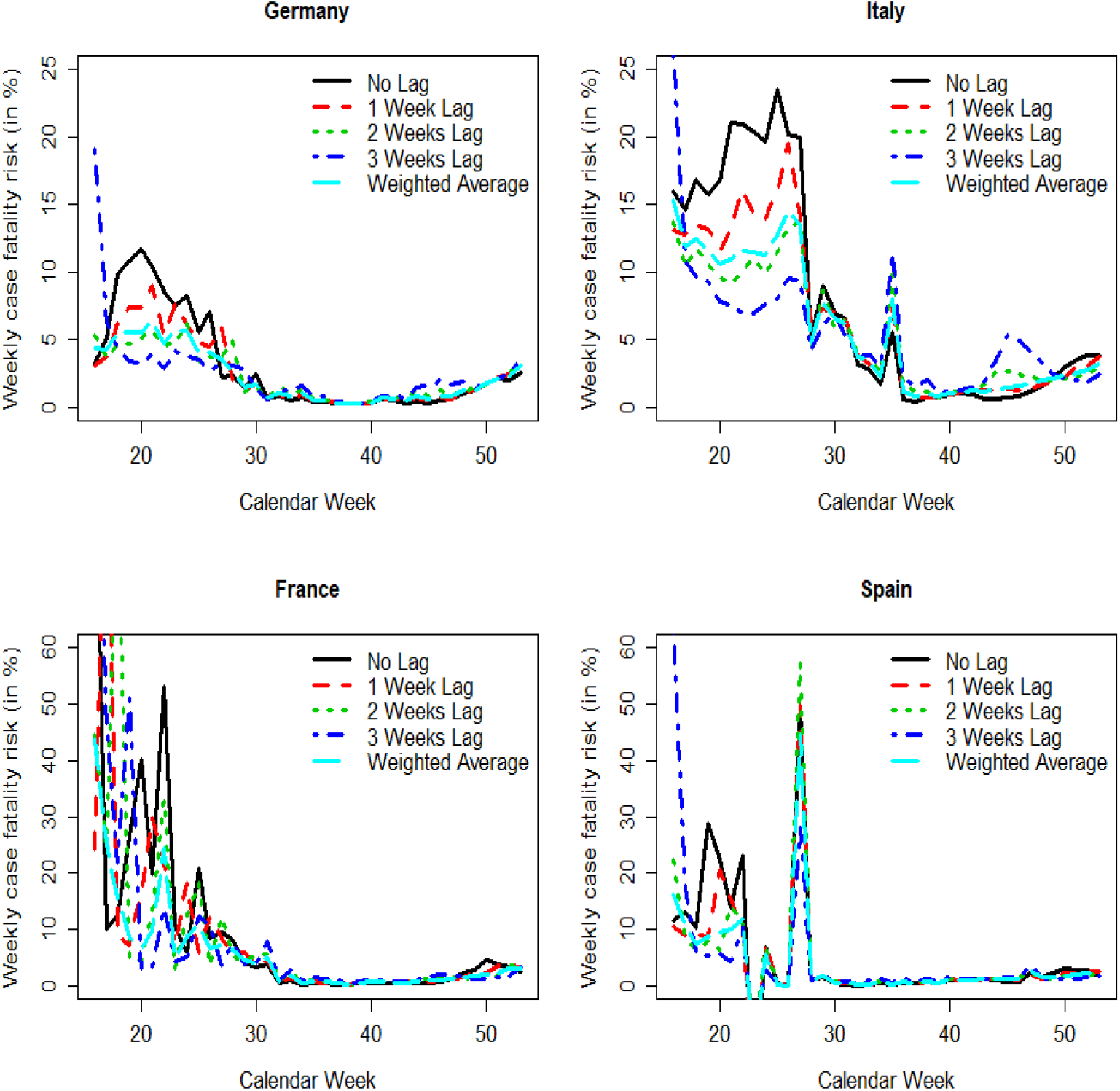
Weekly CFRs with lags 0, 1, 2 or 3 weeks of cases. Sources: Johns Hopkins University (2021); Own computation and design

#### Aim 3. Assess different potential time lags for the CFR estimates

For this part of the analysis, we graphically consider the weekly CFR estimates for the four study countries. Next to the weekly CFR without any lags of case numbers, the CFR series are computed for case lags of one, two and three weeks. Additionally, we include a weighted average of the case numbers as suggested in (9).

All country-specific curves in tendency converge over time and appear to overestimate the CFR at the early stage of the pandemic. The curves which consider lags of higher order appear to perform rather poorly at the beginning but seem to approximate the CFR better after a “burn-in” phase of several weeks. As the time to death varies to some degree by the individual, a weighted average of the cases should smooth strong fluctuations in the case series a bit and might be able to participate in the respective advantages between using no or just one lag and bigger lags of two to three weeks. Graphically, this seems to be the case, as the fluctuations in the weighted average series appear more subtle. This is backed by considering the variance of the series. Among the five alternatives, the weighted average series for Germany, France and Spain has a smaller variance than all of the four series which use only one lag of 0-3 weeks of the case numbers as the denominator. Only for Italy, a lag of 3 weeks causes a smaller variance than the weighted average (0.0021 against 0.0024). Therefore, a weighted lag as in (13) for the cases is the overall best alternative for CFR estimation.

### Aim 4. Describe the association between case fatality and the demand for and supply with hospital beds

Figure 5 illustrates scatterplots of the weekly CFRs, regardless of country ^1^ against 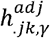 over the study period without any lags, with lags of no and one week, the previous two weeks and the previous three weeks, respectively.

**Figure 5.**
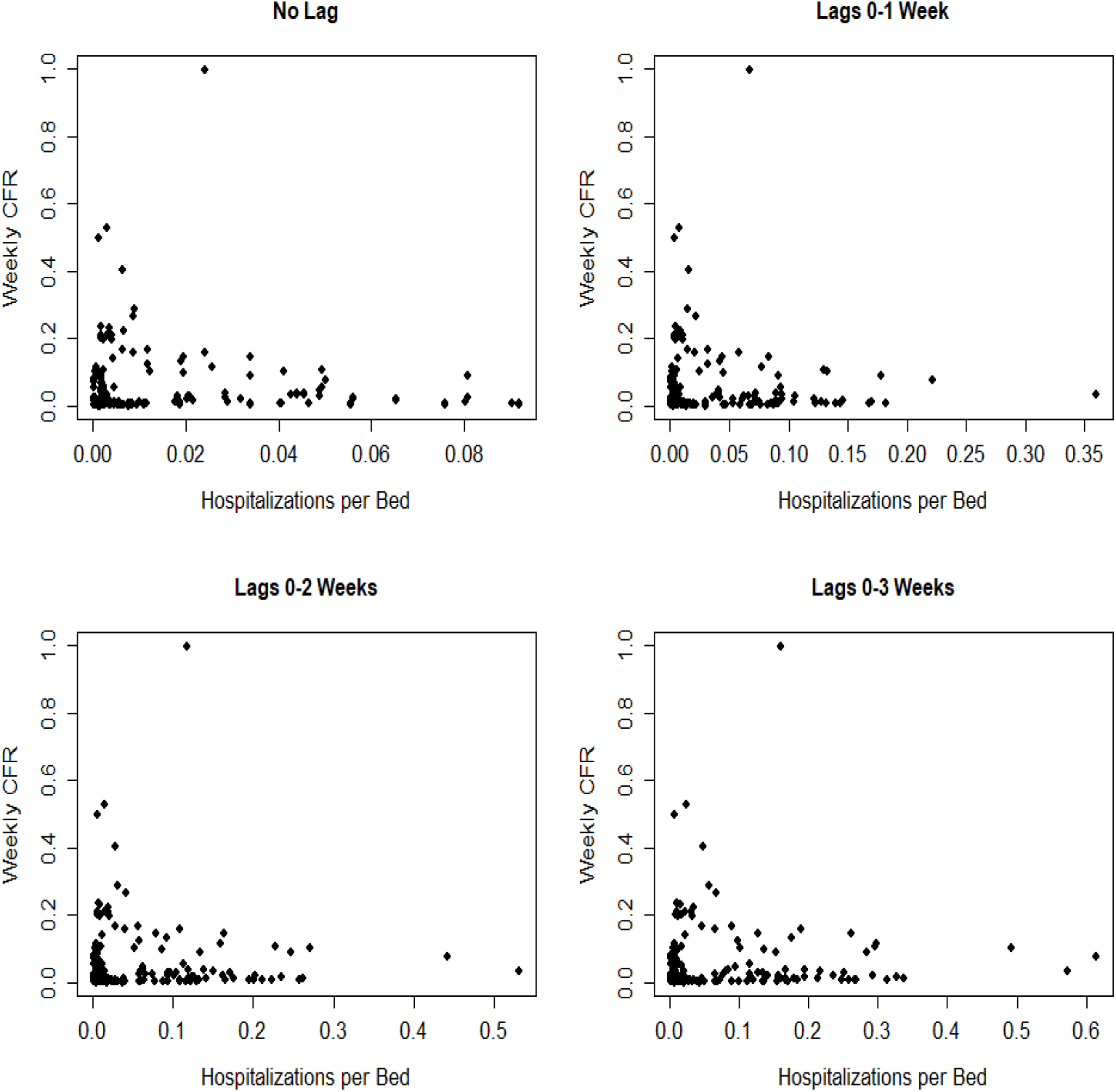
Weekly CFRs against hospitalisations per hospital bed with different lags. Sources: Johns Hopkins University (2021); European Centre for Disease Prevention and Control (2021a); OECD (2021b); Own computation and design

Regardless of which lags are chosen for the COVID-19-related hospitalisations, the weekly CFRs do not show any statistical correlation of our hospital burden to the COVID-19-specific fatality rates.

We will discuss our results in the next section.

## Discussion and conclusion

There are large differences in the reported CFRs between countries. We discussed factors that may explain shares of these differences. We present evidence that a large proportion of the differences of CFRs between the four countries investigated here can be attributed to different age distributions of cases and testing policy. Moreover, we have shown, that crude CFR estimates are strongly biased at peak phases of the pandemic due to lags between case reporting and death. This bias becomes smaller when daily case numbers decrease. Computing CFRs based on weighted averages of the case numbers has a smoothing effect on the CFRs. For the countries investigated, we did not find any connection between healthcare capacity and CFRs of COVID-19.

However, there are still differences in CFRs among the countries, which could not be explained by the factors investigated here. Future research could address those in more detail. We have to keep in mind, however, that there are general differences in age-specific mortality rates between the countries investigated here (Vanella 2017). A more thorough comparative international analysis might take differences in general mortality into account as well. There are certainly other factors, which play into country-specific differences. Among those may be environmental factors, such as air pollution or climatic circumstances (Contini and Costabile 2020). A limitation of our contribution lies in the unobservability of infections. We do not know the real number of infections in the population. To account for this, we included the weekly test rates under assumptions on the relation between testing and detection rates of cases. However, this connection was based on theoretical considerations and does not necessarily hold. A significant limitation of our study is the separate inspection of the impact of the four discussed factors on the CFR estimates. A holistic analysis should consider the joint impact of demographics, detection rates, and time lags on the CFR estimates in a quantitative analysis. We decided against this, as there were no reliable age-specific time series data on detection rates or age-specific test rates available for our study countries over the study period. Therefore, our analysis remains rather qualitative but does not deliver “true” CFRs.

As another important limitation of our work, we found that public data on the age structure of infected and deceased were missing in public reports on COVID-19 in many European countries. Even for the included countries, this data was partly only available at specific time points, for roughly aggregated age groups or only for a selection of all reported cases or deaths. For other countries, age-specific data are not publicly available at all. Moreover, many countries do not even provide data stratified by sex. This biases our understanding of the severity of the disease, as genders show significant differences in susceptibility to severe disease and general mortality (Spagnolo, Manson, and Joffe 2020; Vanella 2017; Luy and Di Giulio 2006). For the analysis of the association between fatality and the healthcare load measured by hospitalisations due to COVID-19, we could not incorporate the age structure or severity of hospitalised cases into our computations, because these data were not available.

Therefore, our study shows the need for further improvements in age-specific data on cases, deaths, and test rates to allow a more sophisticated statistical analysis. For publicly available data to have public health consequences, better reporting of data on healthcare capacities on a daily or at least weekly scale is needed. More detailed data on the demographics of the cases and deaths and age-specific test data would help our understanding of the demographic impact on the CFRs. Even health authorities offering data on the age structure of the cases and deaths do not separate the age groups in the same manner. Important databases give only the crude case and death numbers, without further disaggregation, which might lead to misinterpretation of the true mortality differences among the countries. Moreover, this data should be merged with comorbidity-specific information.

Our study has shown that further improvement towards a better coordinated and unified public health data reporting system in Europe and worldwide is highly warranted to fight this and any other pandemic that may emerge in the future.

## Supporting information

Supplement: Age-standardized CFRs

## Data Availability

Data is available on reasonable request.

## List of abbreviations

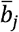: Overall available hospital beds per 1,000 inhabitants in country j
CFR: Case fatality risk
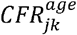: Age-standardised case fatality risk for country j, in week k
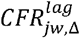: Lag-adjusted case fatality risk for country j, in week w, with *Δ* weekly lags
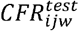: Test-standardised case fatality risk for age group i, in country j, up to week w
*d*_ijk_: Number of deaths in age group i, in country j, in week k
*d*_.jk_: Number of deaths over all age groups, in country j, in week k
*D*_ijw_: Cumulative number of deaths for age group i, in country j, up to week w
*D*_.jw_: Cumulative number of deaths over all age groups, in country j, up to week w
*Δ*: Lag length
ECDC: European Centre for Disease Prevention and Control
EU: European Union
_jk,y_: number of hospitalisations per 100,000 inhabitants in country j, from weeks k-*γ* to k
i.e.: id est
JHU: Johns Hopkins University
*max*{*t*}: Global maximum of weekly tests per 100,000 inhabitants
*n*_ijk_: Number of infections in age group i, in country j, in week k
*N*_ijk_: Cumulative number of infections in age group i, in country j, up to week w
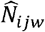: Cumulative number of observed cases for age group i, country j, up to week w
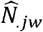: Cumulative number of observed cases for over all age groups, country j, up to week w
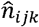: Number of observed cases in age group i, in country j, in week k
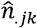: Number of observed cases over all age groups, in country j, in week k
*Ñ*_ijw_: Cumulative test-adjusted number of cases in age group i, in country j, up to week w
*ñ*_ijk_: Test-adjusted number of cases in age group i, in country j, in week k
*ñ*^*^_ijk_: Lag-weighted and test-adjusted number of cases in age group i, in country j, in week k
OECD: Organisation for Economic Co-operation and Development
*t*_jk_: Rate of tests per 100,000 inhabitants in country j, in week k
*w*_i_: Weight of age group i

## Funding

PV, BL and GK received funding from the European Union’s Horizon 2020 research and innovation program under grant agreement No 101003480 and from the Initiative and Networking Fund of the Helmholtz Association.

## Authors’ contributions

HB, PV, CW and BL developed the methods used in this study. PV, HB and CW researched, organised and structured the data and conducted the computations. BL, AM, AH, SW, HB and PV did the literature research. PV built the illustrations of the manuscript. All authors wrote and revised the text.

## Acknowledgments

We thank Gabriele Doblhammer, Carlo-Giovanni “Giancarlo” Camarda and Yohann “Jon” Anson for helpful comments on an earlier version of the paper, which contributed to significant improvements.

## Footnotes

1 We checked this by country as well but the outcome did not change significantly when inspecting individual countries.

## Notes

### Competing Interest Statement

The authors have declared no competing interest.

### Summary of Updates

This paper has been updated until the end of year 2020. Moreover, the analysis has been enhanced.

## References

Cai, H. 2020. “Sex difference and smoking predisposition in patients with COVID-19.” The Lancet Respiratory Medicine 8 (4): E20. https://doi.org/10.1016/s2213-2600(20)30117-x.

Contini, D., and F. Costabile. 2020. “Does Air Pollution Influence COVID-19 Outbreaks?” Atmosphere 11 (4): 377.

Dudel, C., T. Riffe, E. Acosta, A. van Raalte, C. Strozza, and M. Myrskylä. 2020. “Monitoring trends and differences in COVID-19 case-fatality rates using decomposition methods: Contributions of age structure and age-specific fatality.” PLoS ONE 15 (9): e0238904. https://doi.org/10.1371/journal.pone.0238904.

Eriksson, C.O., R.C. Stoner, K.B. Eden, C.D. Newgard, and J.M. Guise. 2017. “The Association Between Hospital Capacity Strain and Inpatient Outcomes in Highly Developed Countries: A Systematic Review.” J Gen Intern Med 32 (6): 686–696. https://doi.org/10.1007/s11606-016-3936-3. https://doi.org/10.1007/s11606-016-3936-3.

European Centre for Disease Prevention and Control. 2020a. Novel coronavirus disease 2019 (COVID-19) pandemic: increased transmission in the EU / EEA and the UK - sixth update. 2020 Mar 12 [cited 2020 Apr 18]. https://www.ecdc.europa.eu/sites/default/files/documents/RRA-sixth-update-Outbreak-of-novel-coronavirus-disease-2019-COVID-19.pdf. https://www.ecdc.europa.eu/sites/default/files/documents/RRA-sixth-update-Outbreak-of-novel-coronavirus-disease-2019-COVID-19.pdf.

Eriksson, C.O. 2020b. “Sentinel surveillance.” Last Modified 2020 Mar 24. Accessed 2020 Mar 24. https://www.ecdc.europa.eu/en/seasonal-influenza/surveillance-and-disease-data/facts-sentinel-surveillance.

Eriksson, C.O. 2021a. “Data on hospital and ICU admission rates and current occupancy for COVID-19.” ECDC. Last Modified 01 March 2021. Accessed 01 March 2021. https://www.ecdc.europa.eu/en/publications-data/download-data-hospital-and-icu-admission-rates-and-current-occupancy-covid-19.

Eriksson, C.O. 2021b. Data on testing for COVID-19 by week and country. European Centre for Disease Prevention and Control, (Solna: European Centre for Disease Prevention and Control). https://www.ecdc.europa.eu/en/publications-data/covid-19-testing.

Eurostat. 2019. “Eurostat Statistics Explained: Healthcare resources statistics - beds.” Last Modified 2019 Nov 29. Accessed 2020 Apr 18. https://ec.europa.eu/eurostat/statistics-explained/index.php/Healthcare_resource_statistics_-_beds.

Eriksson, C.O. 2020. “Population on 1 January by age and sex.” Last Modified 2020 Apr 18. Accessed 2020 Apr 18. https://ec.europa.eu/eurostat/data/database.

Fang, Y., H. Zhang, J. Xie, M. Lin, L. Ying, P. Pang, and W. Ji. 2020. “Sensitivity of Chest CT for COVID-19: Comparison to RT-PCR.” Radiology: 200432. https://doi.org/10.1148/radiol.2020200432.

Gesundheitsberichtserstattung des Bundes. 17 April 2020 2020. Im Informationssystem der GBE zur Altersstandardisierung benutzte Standardbevölkerungen. Gliederungsmerkmale: Alter, Geschlecht, Standardbevölkerung. Destatis, Robert Koch Institut (Berlin, Wiesbaden). http://www.gbe-bund.de/.

Gianicolo, E., N. Riccetti, M. Blettner, and A. Karch. 2020. “Epidemiological Measures in the Context of the COVID-19 Pandemic.” Deutsches Ärzteblatt International 117: 336–342. https://doi.org/10.3238/arztebl.2020.0336.

Gordis, L. 2014. Epidemiology. 5th ed. Philadelphia: Elsevier Saunders. Instituto de Salud Carlos III. 2020a. Informe sobre la situación de COVID-19 en España. Informe COVID-19 no 6-32. 05 de marzo - 21 de mayo de 2020. (Madrid). https://www.isciii.es/QueHacemos/Servicios/VigilanciaSaludPublicaRENAVE/EnfermedadesTransmisibles/Paginas/InformesCOVID-19.aspx.

Gordis, L. 2020b. Informes no 34-59. Situación de COVID-19 en España. Casos diagnosticados a partir 10 de mayo. Informe COVID-19. 15 de julio - 29 de diciembre de 2020. Instituto de Salud Carlos III (Madrid: Instituto de Salud Carlos III). https://www.isciii.es/QueHacemos/Servicios/VigilanciaSaludPublicaRENAVE/EnfermedadesTransmisibles/Paginas/InformesCOVID-19.aspx.

Istituto Superiore di Sanità. 19 March - 29 December 2020 2020a. Epidemia COVID-19. Aggiornamenti nazionali. 19 marzo - 29 dicembre 2020. Istituto Superiore di Sanità, (Roma). https://www.epicentro.iss.it/coronavirus/sars-cov-2-sorveglianza-dati.

Gordis, L. 2020b. “Letalità in Italia minore di quella della Cina per tutte le fasce di età. Comunicati stampa 16/2020.” Last Modified 2020 Mar 06. Accessed 2020 Apr 01. https://www.iss.it/en/comunicati-stampa/-/asset_publisher/fjTKmjJgSgdK/content/id/5288119?_com_liferay_asset_publisher_web_portlet_AssetPublisherPortlet_INSTANCE_fjTKmjJgSgdK_redirect=https%3A%2F%2Fwww.iss.it%2Fen%2Fcomunicati-stampa%3Fp_p_id%3Dcom_liferay_asset_publisher_web_portlet_AssetPublisherPortlet_INSTANCE_fjTKmjJgSgdK%26p_p_lifecycle%3D0%26p_p_state%3Dnormal%26p_p_mode%3Dview%26_com_liferay_asset_publisher_web_portlet_AssetPublisherPortlet_INSTANCE_fjTKmjJgSgdK_cur%3D0%26p_r_p_resetCur%3Dfalse%26_com_liferay_asset_publisher_web_portlet_AssetPublisherPortlet_INSTANCE_fjTKmjJgSgdK_assetEntryId%3D5288119.

Gordis, L. 2020c. “Sorveglianza Integrata COVID-19 in Italia. AGGIORNAMENTO 11 marzo 2020.” Last Modified 2020 Mar 11. Accessed 2020 Mar 12. https://www.epicentro.iss.it/coronavirus/sars-cov-2-sorveglianza-dati.

Gordis, L. 2020d. “Studio ISS Su 105 deceduti con Covid-2019, età media 81 anni e patologie preesistenti in due terzi dei casi. Comunicati stampa 15/2020.” Last Modified 2020 Mar 05. Accessed 2020 Apr 01. https://www.iss.it/en/comunicati-stampa/-/asset_publisher/fjTKmjJgSgdK/content/id/5286166?_com_liferay_asset_publisher_web_portlet_AssetPublisherPortlet_INSTANCE_fjTKmjJgSgdK_redirect=https%3A%2F%2Fwww.iss.it%2Fen%2Fcomunicati-stampa%3Fp_p_id%3Dcom_liferay_asset_publisher_web_portlet_AssetPublisherPortlet_INSTANCE_fjTKmjJgSgdK%26p_p_lifecycle%3D0%26p_p_state%3Dnormal%26p_p_mode%3Dview%26_com_liferay_asset_publisher_web_portlet_AssetPublisherPortlet_INSTANCE_fjTKmjJgSgdK_cur%3D0%26p_r_p_resetCur%3Dfalse%26_com_liferay_asset_publisher_web_portlet_AssetPublisherPortlet_INSTANCE_fjTKmjJgSgdK_assetEntryId%3D5286166.

Ji, Y., Z. Ma, M.P. Peppelenbosch, and Q. Pan. 2020. “Potential association between COVID-19 mortality and health-care resource availability.” The Lancet Glob Health 8 (4): e480. https://doi.org/https://doi.org/10.1016/S2214-109X(20)30068-1. http://www.sciencedirect.com/science/article/pii/S2214109X20300681.

Johns Hopkins University. 2021. “COVID-19 Dashboard by the Center for Systems Science and Engineering (CSSE) at Johns Hopkins University (JHU).” Johns Hopkins University. Last Modified 15 January 2021. Accessed 15 January 2021. https://gisanddata.maps.arcgis.com/apps/opsdashboard/index.html#/bda7594740fd40299423467b48e9ecf6.

Jordan, R.E., P. Adab, and K.K. Cheng. 2020. “Covid-19: risk factors for severe disease and death - A long list is emerging from largely unadjusted analyses, with age near the top.” BMJ 368: m1198. https://doi.org/10.1136/bmj.m1198.

Lau, H., V. Khosrawipour, P. Kocbach, A. Mikolajczyk, H. Ichii, J. Schubert, J. Bania, and T. Khosrawipour. 2020. “Internationally lost COVID-19 cases.” J Microbiol Immunol Infect S1684-1182(20)30073-6. https://doi.org/10.1016/j.jmii.2020.03.013.

Legido-Quigley, H., N. Asgari, Y.Y. Teo, G.M. Leung, H. Oshitani, K. Fukuda, A.R. Cook, L.Y. Hsu, K. Shibuya, and D. Heymann. 2020. “Are high-performing health systems resilient against the COVID-19 epidemic?” The Lancet 395 (10227): 848–850. https://doi.org/https://doi.org/10.1016/S0140-6736(20)30551-1. http://www.sciencedirect.com/science/article/pii/S0140673620305511.

Li, R., S. Pei, B. Chen, Y. Song, T. Zhang, W. Yang, and J. Shaman. 2020. “Substantial undocumented infection facilitates the rapid dissemination of novel coronavirus (COVID-19).” Science 368 (6490): 489–493. https://doi.org/10.1101/2020.02.14.20023127. https://www.medrxiv.org/content/10.1101/2020.02.14.20023127v1.full.pdf.

Lipsitch, M. 2020. “Correspondence to: Estimating case fatality rates of COVID-19.” Lancet Infect Dis doi: 10.1016/S1473-3099(20)30245-0. https://doi.org/10.1016/S1473-3099(20)30245-0.https://www.ncbi.nlm.nih.gov/pubmed/32243813

Lipsitch, M., C.A. Donnelly, C. Fraser, I. M. Blake, A. Cori, I. Dorigatti, N.M. Ferguson, T. Garske, H.L. Mills, S. Riley, M.D. Van Kerkhove, and M.A. Hernan. 2015. “Potential Biases in Estimating Absolute and Relative Case-Fatality Risks during Outbreaks.” PLoS Negl Trop Dis 9 (7): e0003846. https://doi.org/10.1371/journal.pntd.0003846. https://www.ncbi.nlm.nih.gov/pubmed/26181387.

., and P. Di Giulio. 2006. “The Impact of Health Behaviors and Life Quality on Gender Differences in Mortality.” In Gender und Lebenserwartung, Gender kompetent - Beiträge aus dem GenderKompetenzZentrum, edited by J.; Kühl Geppert, J., 113–147. Bielefeld: Kleine.

Ministère des Solidarités et de la Santé. 2021. Données relatives à l’épidémie de COVID-19 en France : vue d’ensemble. Ministère des Solidarités et de la Santé (Paris). https://www.data.gouv.fr/en/datasets/donnees-relatives-a-lepidemie-de-covid-19-en-france-vue-densemble/#_.

OECD. 15 January 2021 2021a. Beyond Containment: Health systems responses to COVID-19 in the OECD. OECD (Paris: OECD). https://read.oecd-ilibrary.org/view/?ref=119_119689-ud5comtf84&Title=Beyond%20Containment:Health%20systems%20responses%20to%20COVID-19%20in%20the%20OECD.

Luy, M. 2021b. “Hospital beds (indicator).” OECD. Accessed 01 March 2021. https://data.oecd.org/healtheqt/hospital-beds.htm.

Onder, G., G. Rezza, and S. Brusaferro. 2020. “Case-Fatality Rate and Characteristics of Patients Dying in Relation to COVID-19 in Italy.” JAMA doi: 10.1001/jama.2020.4683. https://doi.org/10.1001/jama.2020.4683. https://www.ncbi.nlm.nih.gov/pubmed/32203977

https://jamanetwork.com/journals/jama/articlepdf/2763667/jama_onder_2020_vp_200059.pdf.

Pan, Y., D. Zhang, P. Yang, L.L.M. Poon, and Q. Wang. 2020. “Viral load of SARS-CoV-2 in clinical samples.” The Lancet Infect Dis 20 (4): 411–412. https://doi.org/10.1016/s1473-3099(20)30113-4.

Rajgor, D.D., M.H. Lee, S. Archuleta, N. Bagdasarian, and S.C. Quek. 2020. “The many estimates of the COVID-19 case fatality rate.” The Lancet Infect Dis doi: 10.1016/S1473-3099(20)30244-9. https://doi.org/10.1016/S1473-3099(20)30244-9. https://doi.org/10.1016/S1473-3099(20)30244-9.

Reich, N.G., J. Lessler, D.A.T. Cummings, and R. Brookmeyer. 2012. “Estimating Absolute and Relative Case Fatality Ratios from Infectious Disease Surveillance Data.” Biometrics 68 (2): 598–606.

Rhodes, A., P. Ferdinande, H. Flaatten, B. Guidet, P.G. Metnitz, and R.P. Moreno. 2012. “The variability of critical care bed numbers in Europe.” Intensive Care Med 38 (10): 1647–1653. https://doi.org/10.1007/s00134-012-2627-8. https://doi.org/10.1007/s00134-012-2627-8.

Robert Koch Institut. 2020a. SARS-CoV-2-Testkriterien für Schulen während der COVID-19-Pandemie. Empfehlungen des Robert Koch-Instituts für Schulen. Robert Koch Institut (Berlin: Robert Koch Institut). https://www.rki.de/DE/Content/InfAZ/N/Neuartiges_Coronavirus/Teststrategie/Nat-Teststrat.html.

Rhodes, A. 2020b. “Syphilis in Deutschland 2019 – Neuer Höchststand von Infektionen; SARS-CoV-2-Testzahlen in Deutschland.” Epidemiologisches Bulletin 2020 (49). https://www.rki.de/DE/Content/Infekt/EpidBull/Archiv/2020/Ausgaben/49_20.pdf?__blob=publicationFile.

Rhodes, A. 2021. RKI COVID19. Robert Koch Institut, (Berlin: Robert Koch Institut). https://www.arcgis.com/home/item.html?id=f10774f1c63e40168479a1feb6c7ca74.

Santé publique France. 2020a. “COVID-19: Point épidémiologique - Situation au 4-15 mars 2020 à minuit.” Last Modified 2020 Mar 15. Accessed 2020 Mar 20. https://www.santepubliquefrance.fr/recherche/#search=COVID-19%20:%20point%20%C3%A9pid%C3%A9miologique.

Rhodes, A. 2020b. COVID-19: Point épidémiologique hebdomaire du 24 mars - 31 décembre 2020. Santé publique France, (Saint-Maurice). https://www.santepubliquefrance.fr/.

Shim, E., A. Tariq, W. Choi, Y. Lee, and G. Chowell. 2020. “Transmission potential and severity of COVID-19 in South Korea.” Int J Infect Dis 93: 339–344. https://doi.org/https://doi.org/10.1016/j.ijid.2020.03.031. http://www.sciencedirect.com/science/article/pii/S1201971220301508.

Spagnolo, P.A., J.E. Manson, and H. Joffe. 2020. “Sex and Gender Differences in Health: What the COVID-19 Pandemic Can Teach Us.” Annals of Internal Medicine doi: 10.7326/M20-1941.

Spychalski, P., A. Blazynska-Spychalska, and J. Kobiela. 2020. “Correspondence to: Estimating case fatality rates of COVID-19.” Lancet Infect Dis doi: 10.1016/S1473-3099(20)30246-2. https://doi.org/10.1016/S1473-3099(20)30246-2. https://www.ncbi.nlm.nih.gov/pubmed/32243815.

Vanella, P. 2017. “A principal component model for forecasting age-and sex-specific survival probabilities in Western Europe.” Zeitschrift für die gesamte Versicherungswissenschaft (German Journal of Risk and Insurance) 106 (5): 539–554.

Verity, R., L.C. Okell, I. Dorigatti, P. Winskill, C. Whittaker, N. Imai, G. Cuomo-Dannenburg, H. Thompson, P.G.T. Walker, H. Fu, A. Dighe, J.T. Griffin, M. Baguelin, S. Bhatia, A. Boonyasiri, A. Cori, Z. Cucunubá, R. FitzJohn, K. Gaythorpe, W. Green, A. Hamlet, W. Hinsley, D. Laydon, G. Nedjati-Gilani, S. Riley, S. van Elsland, E. Volz, H. Wang, Y. Wang, X. Xi, C.A. Donnelly, A.C. Ghani, and N.M. Ferguson. 2020. “Estimates of the severity of coronavirus disease 2019: a model-based analysis.” Lancet Infect Dis doi: 10.1016/S1473-3099(20)30243-7.

Wilson, N., A. Kvalsvig, L. T. Barnard, and M. G. Baker. 2020. “Case-Fatality Risk Estimates for COVID-19 Calculated by Using a Lag Time for Fatality.” Emerg Infect Dis 26 (6). https://doi.org/10.3201/eid2606.200320. https://www.ncbi.nlm.nih.gov/pubmed/32168463.

World Health Organization. 2020a. “Laboratory testing for 2019 novel coronavirus (2019-nCoV) in suspected human cases. Interim guidance.” Last Modified 2020 Mar 19. Accessed 2020 Apr18. https://www.who.int/publications-detail/laboratory-testing-for-2019-novel-coronavirus-in-suspected-human-cases-20200117.

Wilson, N. 2020b. Assessing National Capacity for the Prevention and Control and Noncommunicable Diseases: Report of the 2019 Global Survey. Geneva: World Health Organization.

Wu, J.T., K. Leung, M. Bushman, N. Kishore, R. Niehus, P.M. de Salazar, B.J. Cowling, M. Lipsitch, and G.M. Leung. 2020. “Estimating clinical severity of COVID-19 from the transmission dynamics in Wuhan, China.” Nat Med 26: 506–510. https://doi.org/10.1038/s41591-020-0822-7. https://doi.org/10.1038/s41591-020-0822-7.

Wu, Z., and J.M. McGoogan. 2020. “Characteristics of and Important Lessons From the Coronavirus Disease 2019 (COVID-19). Summary of a Report of 721314 Cases From the Chinese Center for Disease Control and Prevention.” JAMA 323 (13): 1239–1242.

Xie, J., Z. Tong, X. Guan, B. Du, H. Qiu, and A.S. Slutsky. 2020. “Critical care crisis and some recommendations during the COVID-19 epidemic in China.” Intensive Care Med 46 (5): 837–840. https://doi.org/10.1007/s00134-020-05979-7.

Yang, J., Y. Zheng, X. Gou, K. Pu, Z. Chen, Q. Guo, R. Ji, H. Wang, Y. Wang, and Y. Zhou. 2020. “Prevalence of comorbidities in the novel Wuhan coronavirus (COVID-19) infection: a systematic review and meta-analysis.” Int J Infect Dis 94: 91–95. https://doi.org/10.1016/j.ijid.2020.03.017.

Zhang, J.J., X. Dong, Y.Y. Cao, Y.D. Yuan, Y.B. Yang, Y.Q. Yan, C.A. Akdis, and Y.D. Gao. 2020. “Clinical characteristics of 140 patients infected with SARS-CoV-2 in Wuhan, China.” Allergy doi: 10.1111/all.14238. https://doi.org/10.1111/all.14238.

Zhou, F., T. Yu, R. Du, G. Fan, Y. Liu, Z. Liu, J. Xiang, Y. Wang, B. Song, X. Gu, L. Guan, Y. Wei, H. Li, X. Wu, J. Xu, S. Tu, Y. Zhang, H. Chen, and B. Cao. 2020. “Clinical course and risk factors for mortality of adult inpatients with COVID-19 in Wuhan, China: a retrospective cohort study.” Lancet 395 (10229): 1054–1062. https://doi.org/10.1016/S0140-6736(20)30566-3.

